## Supplemental materials for "An In-Depth Evaluation of Federated Learning on Biomedical Natural Language Processing"

### Supplementary Materials

#### Experiment details

**Data Preprocessing**: We removed duplicated notes and split the data into the non-overlapped train(80%), dev(10%), and test(10%) datasets. To simulate an FL setting, we further split the train and dev set equally into 2/5/10 folds to mimic varied numbers of clients' participation.

**Hyperparameter tuning**: For models involving one or more hyper-parameters, we applied grid search to find the best combination of hyper-parameters.

**FL Algorithms:** Considering there are numerous FL algorithms designed to tackle various problems, we pick two fundamental algorithms, FedAvg and FedProx, both are the most representative and popular FL algorithms that are widely used in practice. We showed the comparisons between the two algorithms as detailed in Algorithm 1 (server aggregation) and Algorithm 2 (clients' local updates).

---

**Algorithm 1**: Federated learning algorithms (FedAvg/FedProx)

---

Notation: $X_i$ indicate data from client i, K is the total number of clients, T is maximum training round, n is the sum of $n_1$ to $n_k$, σ is the hyper-parameter in FedAMP

Initialize server model weights $\mathbf{w}(1)$
Initialize client model weights $w_i\ \forall\ i = 1, 2, ..., K$
For each round t = 1, 2, … T do
    Send server model weight $w(t)$ to each client
    For each client $k = 1, 2, ..., K$ do
        Client k perform LocalUpdate($X_k$, $Y_k$, $w_k$)     ← Algorithm 2

$\gamma_k = \frac{n_k}{n}$

End for

End for

**Algorithm 2**: Local model training using mini-batch stochastic gradient descent (LocalUpdate) (FedAvg/FedProx)

Notation: R is the local update round, B is the number of batches, $f_{w_r}$ is the neural network parameterized by $w_r$, η is the learning rate, μ is the hyper-parameter in FedProx, λ and $\alpha_k$ is the hyper-parameters in FedAMP

For each round $r = 1, 2, ..., R$ do / Repeat until find the approximate minimizer of $w \approx argmin_w L(f_{w_r}(X_b), Y_b) + \frac{\mu}{2}||w_k - w_k(t)||^2$

Randomly shuffle $X_k$ and create B batches $((X_1, Y_1), (X_2, Y_2), ..., (X_B, Y_B))$

$L_{w_r} = L(f_{w_r}(X_b), Y_b) + \frac{\mu}{2}||w_k - w_k(t)||^2$

For each mini-batch $b = 1, 2, ..., B$ do

$w_{r+1} = w_r - \eta \nabla L_{w_r}(X_b, Y_b)$

End for

#### Datasets

**2018 National NLP Clinical Challenges (n2c2) Shared Task**[38]: 2018 n2c2 corpus contains 505 discharge summaries from the MIMIC-III clinical care database[4]. The goal of the task is to extract entity tags (*reason, frequency, ADE, strength, duration, route, form, drug, and dosage*) that indicate the presence of drug and ADE information, and relations (*strength-drug, duration-drug, route-drug, form-drug, ADE-drug, Dosage-drug, reason-drug, and frequency-drug*) between the entities.

**BioCreative II Gene Mention Recognition (BC2GM)**[39]: BC2GM Dataset collected text data related to gene information. The dataset comprises a set of sentences, and a set of gene mentions (GENE annotations) for each sentence. Some GENE annotations in a sentence may also have alternate boundaries that are judged by human annotators which can be essentially equivalent references (ALTGENE annotations). The goal of the task is to identify gene mentions in a sentence according to its start and end characters.

**BioCreative IV Chemical Compound and Drug Name Recognition (BC4CHEMD)**[40]: BC4CHEMD contains a total of 84,355 chemical mention annotations from 10,000 PubMed abstracts which are manually labeled by some chemistry literature experts. The goal of the task is to classify the text into multiple CEM classes: *systematic, identifiers, formula, trivial, abbreviation, family, and multiple.*

**JNLPBA**[41]: JNLPBA originated from the GENIA version 3.02. It is a selection of 2,000 abstracts with a controlled search on MEDLINE using the MeSH terms *'human'*, *'blood cells'*, and *'transcription factors'*. The abstracts were hand-annotated to 36 terminal classes according to a small taxonomy of 48 classes based on a chemical classification.

**NCBI-disease**[42]: The NCBI-disease corpus is collected from 793 PubMed abstracts that are fully annotated at the disease mentions and concept level based on corresponding identifiers from either Medical Subject Headings (MeSH) or Online Mendelian Inheritance in Man (OMIM). It includes 6892 disease mentions, which are mapped to 790 unique disease concepts. 12% link to an OMIM identifier, while the remaining contain a MeSH identifier. In addition, 91% of mentions are described as a single disease concept, while the remaining link to a combination of concepts.

**EUADR**[43]: EUADR corpus was annotated for disorders, drugs, genes, and their inter-relationships. Three experts were used to annotate a set of 100 abstracts for each of the drug-disorder, drug-target, and target-disorder relations. The drug-disorder and drug-target relations were composed of 100 randomly selected abstracts from the PubMed result. For the target-disorder set, 50 abstracts were randomly selected from gene disorder, and 50 abstracts were randomly selected from SNP-disorder relation.

**Gene Associations Database (GAD)**[21]: Gene Associations Dataset is a corpus that provides a public, comprehensive repository of molecular, clinical, and study parameters for > 5000 human genetic association studies to explore gene-disease relations. It contains 10697 genes, 12774 diseases, and 74928 gene-disease associations.

#### Models Architectures

**BERT**[1]: Bidirectional Encoder Representations from Transformer (BERT) was developed in 2018 by researchers at Google. The BERT’s model architecture is a multi-layer bidirectional Transformer encoder. It employs encoders as a sub-structure to pre-training models for NLP tasks. BERT comprehends language via Masked Language Modeling (MLM) and Next Sentence Prediction (NSP) mechanisms. By assuming a blinder with MLM, BERT learns bidirectional contexts within sentences. The model takes random sentences as input, masks certain words, and then reconstructs the masked words from the surrounding text. BERT's ability to process two sentences simultaneously and determine if the second follows the first enables it to achieve NSP, facilitating the maintenance of long-distance relationships between texts. The BERT was pre-trained on English Wikipedia (2.5B words) and BookCorpus (800M words). BERT has two models. The BERT_BASE has 12 layers, 768 widths, 12 heads, and a total of 110M parameters. The BERT_LARGE has 24 layers, 1024 widths, 16 heads, and a total of 340M parameters.

**GPT-2**[46]: Generative Pretrained Transformer 2 (GPT2) was developed by OpenAI researchers in 2019. GPT-2 models language using transformer decoders. GPT-2 is designed for predicting the next sentence in sentences. It achieves this using the architecture of GPT-1 with an extra normalization layer to the input of each sub-block and after the final self-attention layer. GPT-2's input includes texts' weight embeddings and their positional embeddings for context extraction. The input is then passed through the multi-head attention layer in the transformer decoder blocks, followed by a feed-forward layer. Finally, the softmax outputs a probability distribution. The GPT-2 was trained on an extensive corpus of WebText (40 GB of text, 8 million documents, from 45 million webpages upvoted on Reddit). It has 48 layers, 1600 widths, and 1.5B parameters.

**BI-LSTM-CRF**[44]: The Bidirectional LSTM CRF network combines a bidirectional LSTM network with a CRF network. It efficiently makes use of both long-distance past and future input features via the bidirectional LSTM layer as well as sentence-level tag information via the CRF layer. The initial layer of the model, designed to capture the semantics of the input text sequence, is a Bi-LSTM network. This layer's output is then fed into a CRF layer, which generates a probability distribution over the tag sequence by utilizing the interdependencies among the labels of the entire sequence.

**GPT-3**[47]: The Generative Pretrained Transformer 3 (GPT-3) employs a similar model architecture as GPT-2; however, it incorporates both alternating dense and locally banded sparse attention patterns in the transformer layers. The latest GPT-3 with 175B parameters was trained on the Common Crawl Dataset, expanded WebText Dataset, two internet-based books corporas, and English Wikipedia, using a context window of 2048 tokens. The pre-training steps mirror those of GPT-2, relatively but with increased dataset scale, data diversity, and training duration. The checkpoint used for experiments in this paper is "gpt-3.5-turbo-0613".

**GPT-4**[48]: The Generative Pretrained Transformer 4 (GPT-4) was released by OpenAI in 2023 without disclosure of model architecture, training, dataset, etc. Different from GPT-3, GPT-4 was fine-tuned using Reinforcement Learning from Human Feedback (RLHF). The checkpoint used for experiments in this paper is GPT-4-0613.

**PaLM 2**[49]: The Pathways Language Model (PaLM) 2 is built upon the Transformer. Specific information regarding its model size and architecture has not been shared in public. From available information, however, the predecessor of PaLM 2, known as PaLM[50], has 540B parameters, and deploys a standard Transformer architecture in a decoder-only configuration, albeit with certain alterations (e.g., SwiGLU Activation, RoPE embedding, etc). The checkpoint used for experiments in this paper is “bison-001”, the second largest model in the PaLM 2 family.
